# A Chronological and Geographical Analysis of Personal Reports of COVID-19 on Twitter

**DOI:** 10.1101/2020.04.19.20069948

**Authors:** Ari Z. Klein, Arjun Magge, Karen O’Connor, Haitao Cai, Davy Weissenbacher, Graciela Gonzalez-Hernandez

**Affiliations:** Department of Biostatistics, Epidemiology, and Informatics, Perelman School of Medicine, University of Pennsylvania, Philadelphia, PA, USA

## Abstract

The rapidly evolving outbreak of COVID-19 presents challenges for actively monitoring its spread. In this study, we assessed a social media mining approach for automatically analyzing the chronological and geographical distribution of users in the United States reporting personal information related to COVID-19 on Twitter. The results suggest that our natural language processing and machine learning framework could help provide an early indication of the spread of COVID-19.

## INTRODUCTION

The rapidly evolving outbreak of COVID-19, and the delay and shortage of available testing in the United States, presents challenges for actively monitoring its spread and preparing in response. One approach for detecting cases without the need of extensive testing relies on voluntary self-reports of symptoms from the general population.^1^ However, the incubation period of COVID-19^2^ may limit active monitoring based primarily on symptoms. Considering that nearly one of every four adults in the United States already uses Twitter, and nearly half of them use it on a daily basis,^3^ in this proof-of-concept study, we assessed (1) whether users report personal information on Twitter that could more broadly indicate potential exposure to COVID-19, and (2) the utility of our social media mining approach for automatically detecting these users and analyzing the chronological and geographical distribution of their reports. To our knowledge, the use of real-time Twitter data to track COVID-19^4^ has not extended to user-level, personal reports. Thus, our natural language processing and machine learning framework could advance the use of Twitter data as a complementary resource “to understand and model the transmission and trajectory of COVID-19”.^5^

To assess whether users report personal information on Twitter that could indicate potential exposure to COVID-19, we manually annotated a random sample of 10,000 pre-filtered tweets, distinguishing three classes:

- *Probable*: The tweet indicates that the user or a member of the user’s household has been diagnosed with, tested for, denied testing for, symptomatic of, or directly exposed to confirmed or presumptive cases of COVID-19.
- *Possible*: The tweet indicates that the user or a member of the user’s household has had experiences that pose a higher risk of exposure to COVID-19 (e.g., recent traveling) or exhibits symptoms that may be, but are less commonly, associated with COVID-19.
- *Other*: The tweet is related to COVID-19 and may discuss topics such as testing, symptoms, traveling, or social distancing, but it does not indicate that the user or a member of the user’s household may be infected.

To demonstrate the utility of the annotated corpus for training machine learning algorithms, we present the benchmark performance of a deep neural network classifier using pre-trained Bidirectional Encoder Representations from Transformers (BERT).^6^ We also present the results of deploying the classifier on unlabeled tweets collected between January 23, 2020 and April 6, 2020, and extracting the date and location—geo-tags or profile metadata—of those that were automatically classified as “probable” or “possible.”

## RESULTS AND DISCUSSION

Evaluated on a held-out test set of 2000 tweets, the BERT-based classifier achieved benchmark F_1_-scores of 0.64 (precision = 0.69, recall = 0.61) for the “probable” class, 0.53 (precision = 0.54, recall = 0.52) for the “possible” class, and 0.68 (precision = 0.70, recall = 0.67) when the “probable” and “possible” classes were unified:

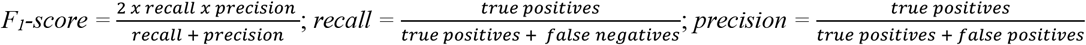

Figure 1 illustrates the number of detected users from U.S. states who have posted “probable” or “possible” tweets between January 23, 2020 and April 6, 2020. Figure 2 illustrates the cumulative number of users from the top 12 states by report date.

**Fig. 1.**
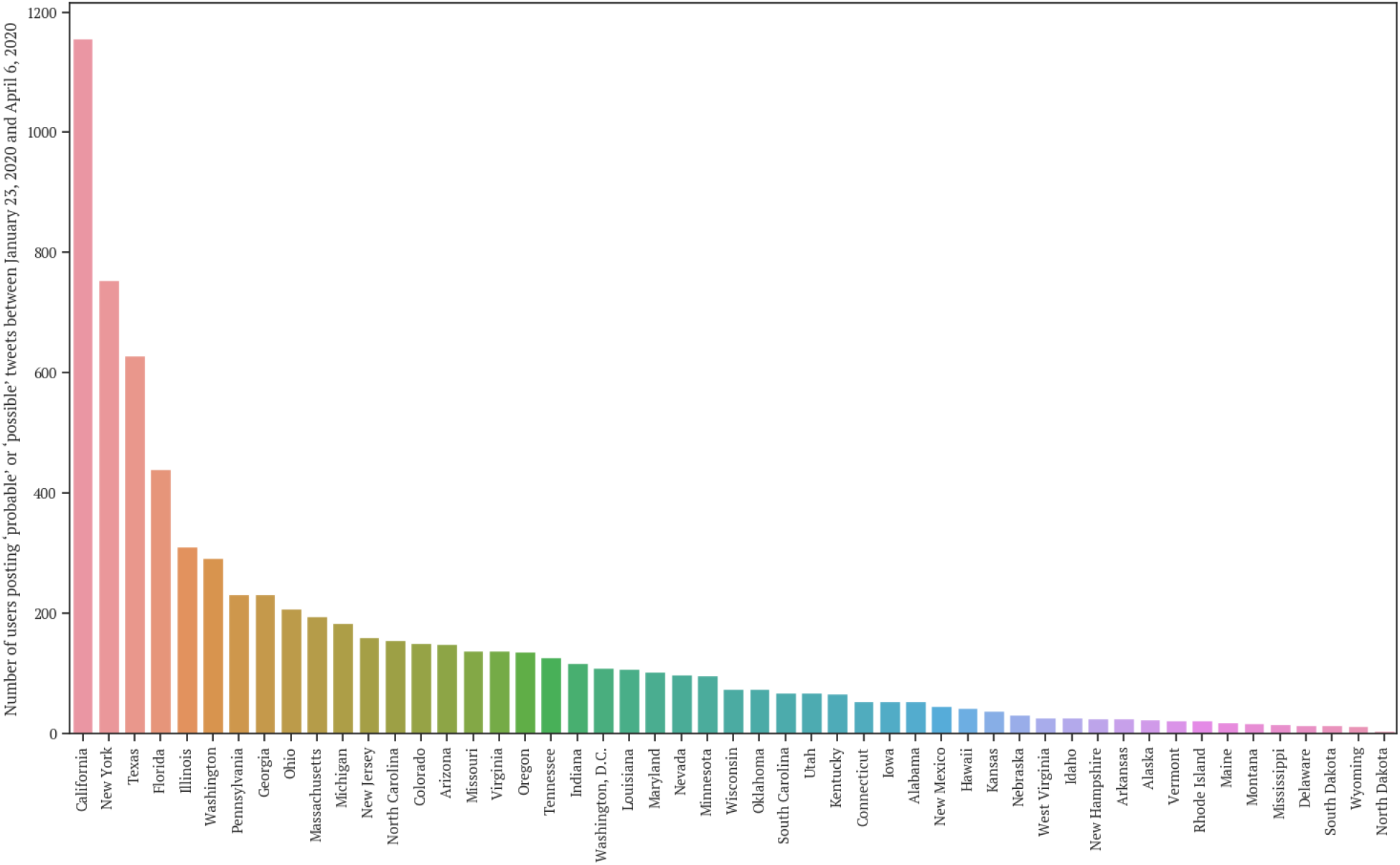
Cumulative number of users posting “probable” or “possible” tweets by state, January 23, 2020 to April 6, 2020

**Fig. 2.**
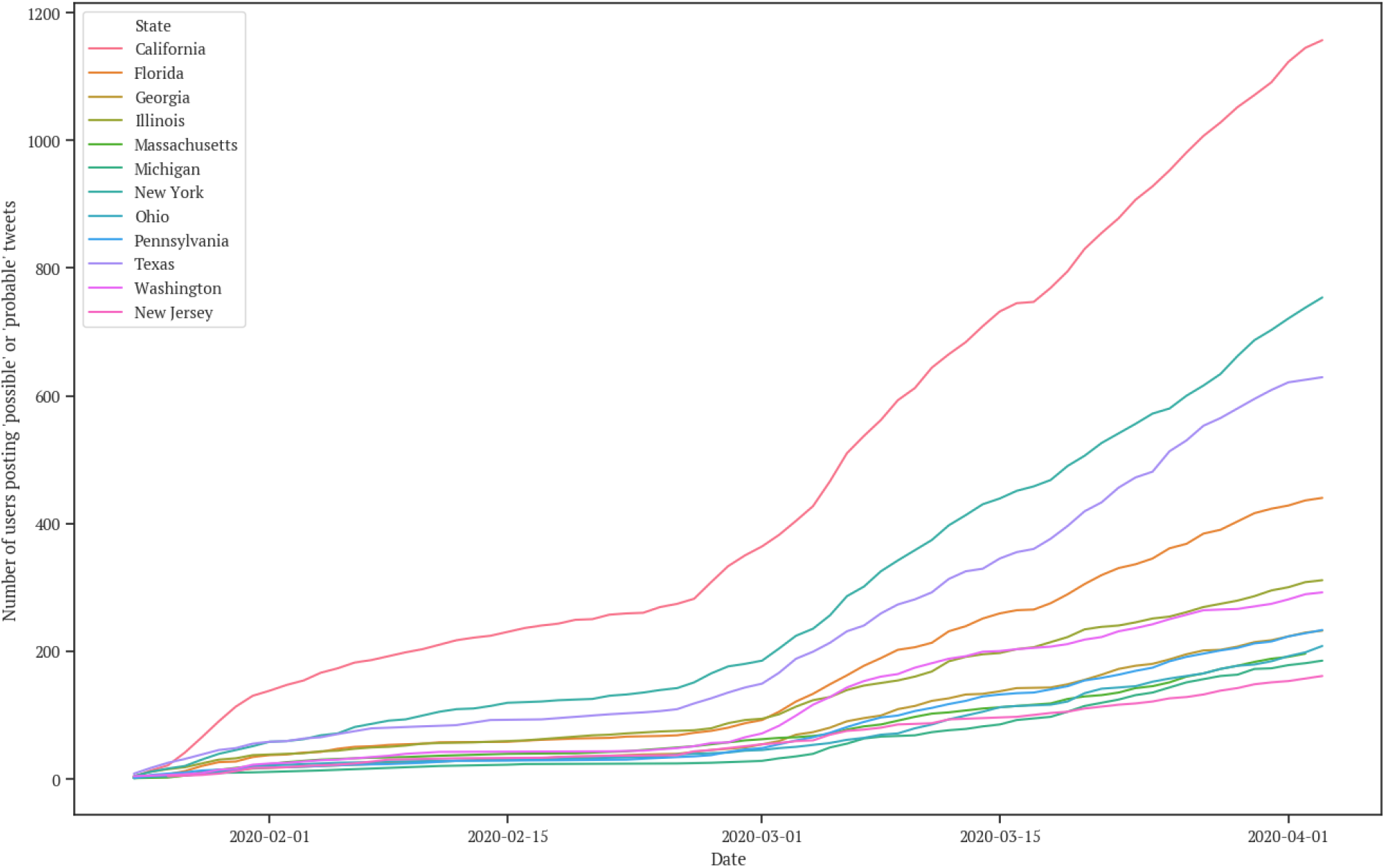
Cumulative number of users from 12 states posting “probable” or “possible” tweets by report date, January 23, 2020 to April 6, 2020

In Figure 1, the 12 states with the most users who have posted “probable” or “possible” tweets include 11 of the top 12 states reporting COVID-19 cases to the CDC^7^. These personal reports on Twitter began to increase sharply around the beginning of March, as shown in Figure 2, but not until the middle/end of March for cases reported to the CDC^7^. For many states, we have detected “probable” or “possible” tweets that were posted before the state’s first confirmed case. Thus, this study demonstrates that (1) users do report personal information on Twitter that could broadly indicate potential exposure to COVID-19, (2) these personal reports can be understood as signals of COVID-19 cases, and (3) our social media mining approach could help provide an early indication of the spread of COVID-19. Despite these promising preliminary results, many of the tweets were not visualized because the locations in the user profile metadata could not be normalized to the state level. We will address this challenge in future work.

## METHODS

### Data Collection and Annotation

This study received an exempt determination by the Institutional Review Board of the University of Pennsylvania, as it does not meet the definition of “human subject” according to 45 CRF § 46.102(f). Between January 23, 2020 and March 20, 2020, we collected more than 7 million publicly available, English tweets (excluding retweets) from the Twitter Streaming API that mention keywords related to COVID-19 and are geo-tagged or have profile location metadata. Then, using handcrafted regular expressions, we identified 160,767 of the tweets that contain information potentially indicating that the user or a member of the user’s household had been exposed to COVID-19. We removed 30,564 of the matching tweets that were automatically determined to contain “reported speech” (e.g., quotations, news headlines) using a filter we developed in recent work. We manually annotated a random sample of 10,000 of the 130,203 pre-filtered tweets. Annotation guidelines (supplemental file) were developed to help two annotators distinguish tweets that indicate (1) a “probable” case of COVID-19, (2) a “possible” case, or (3) merely discuss COVID-19. Inter-annotator agreement was κ = 0.73 (Cohen’s kappa), considered “substantial agreement.”^8^ Upon resolving the disagreements, 6.9% (685) of the 10,000 tweets were annotated as “probable,” 7.8% (780) as “possible,” and 85.3% (8535) as “other.”

### Classification and Normalization

We split the 10,000 annotated tweets into 80% (supplemental file) and 20% random sets to train and evaluate a supervised deep neural network classifier using a pre-trained BERT model with 12 Transformer blocks, 768 units for each hidden layer, and 12 self-attention heads. We used a maximum sequence length of 100 tokens to encode. After feeding the sequence of token IDs to BERT, the encoded representation is passed to a dropout layer (dropping rate of 0.1) and, then, a dense layer with 2 units and a softmax activation, which predicts the class for each tweet. For training, we used Adam optimization with rate decay and warm-up. We used a batch size of 64, training runs for 3 epochs, and a maximum learning rate of 1e-4 for the first 10% of training steps, with the learning rate decaying to 0 in the latter 90% of training steps. Prior to automatic classification, we pre-processed the tweets by normalizing user names (i.e., strings beginning with “@”) and URLs, and lowercasing the text. We deployed the classifier on the 430,574 unlabeled, pre-filtered tweets collected between January 23, 2020 and April 6, 2020. We used GeoNames^9^ to normalize—to the U.S. state level (ADM1)—locations associated with the tweets that were automatically classified as “probable” or “possible.” For tweets without geo-tags, we adapted our previous work^10^ to extract and disambiguate user-generated locations in the profile metadata. We derived one location per user.

## Data Availability

The annotated data that was used to train the classifier for the evaluation in this study is available as a supplemental file with this article. Tweets annotated as “other,” “probable,” and “possible” are labeled as “0,” “1,” and “2,” respectively. To download the tweets, a Python script is available at https://bitbucket.org/pennhlp/twitter_data_download/src/master/.

## ACKNOWLEDGEMENTS

This work was supported by the National Institutes of Health (NIH) National Library of Medicine (NLM) grant R01LM011176 and National Institute of Allergy and Infectious Diseases (NIAID) grant R01AI117011. The content is solely the responsibility of the authors and does not necessarily represent the views of the NIH, NLM, or NIAID. The authors would like to acknowledge Alexis Upshur for contributing to annotating the Twitter data.

## AUTHOR CONTRIBUTIONS

A.Z.K handcrafted the regular expressions, deployed the reported speech filter, prepared the data for annotation, contributed to developing the annotation guidelines, and wrote the article. A.M. extracted the tweet locations and generated the figures. K.O. developed the annotation guidelines and annotated the tweets. H.C. collected the tweets from the Twitter Streaming API and trained, evaluated, and deployed the BERT-based classifier. D.W. performed baseline classification experiments. G.G.H. provided the overall study design. All authors contributed to editing the article.

